# Construction and application of a revised satisfaction index model for Chinese urban and rural residents basic medical insurance

**DOI:** 10.1101/2020.05.23.20108779

**Authors:** Wenwei Cheng, Jin Cheng, Xiaofang Liu, Yanyan Wu, Weichu Sun, Xiaofang Yan, Liai Peng, Xiaoli Liu, Qi Wang, Mingming Luo, Tingting Sha, Jingcheng Shi, Fang Yang

## Abstract

**Background:** Quality is the most important factor of satisfaction However, the existing URRBMI index model lacks the decomposition of the connotation of perceived quality and cannot provide a reference for quality improvement and satisfaction promotion.

**Objective:** This study aims to construct a Satisfaction Index Model for Chinese Urban and Rural Residents Basic Medical Insurance (SIM-URRBMI) with accurate and detailed measurement of perceived quality and give a feasible and scientific suggestion for URRBMI or insurance for other countries in the world.

**Methods:** Based on the theoretical framework of The American Customer Satisfaction Index (ACSI), the connotation of perceived quality was refined by literature review and expert consultation to form a pool of alternative measurement variables. A three-stage randomized stratified cluster sampling was adopted to select the main decision makers for pupils’ URRBMI in 8 primary schools from Changsha City. Both Classic Test Theory (CTT) and Item Response Theory (IRT) were used for selection of the measurement variables. The model’s reliability and validity were tested using partial least square (PLS) related methods.

**Results:** A total of 1909 respondents who had insurance for their children were investigated with the initial questionnaire. The revised SIM-URRBMI consists of 11 latent variables and 28 measurement variables with good reliability and validity. Among the three explanatory variables of public satisfaction, perceived quality had the largest total effect (0.737). The variable with greatest effect among the five first-order latent variables on perceived quality was quality of the medical insurance policy (0.472).

**Conclusions:** The revised SIM-URRBMI consists of 11 latent variables and 28 measurement variables with good reliability and validity. It provides accurate assessment of perceived quality, which will greatly help performance improvement. Perceived quality is crucial to public satisfaction, especially, the most important aspects are policies regarding medical insurance reimbursement (basic coverage scope, coinsurance, deductible).

## Introduction

According to the 2015 World Health Organization (WHO) report, 6% of people in low-and middle-income countries are forced into extreme poverty because of medical expendituresThe Chinese government has been trying to establish a multilevel medical security system that is mainly based on the social basic medical insurance (BMI) schemes so as to protect the finances of individuals and households affected by illness and injury to some extent. In order to improve equity, sustainability, and efficiency, China has taken an initial step to integrate new cooperative medical scheme (NCMS) and urban resident-based basic medical insurance (URBMI) into the urban and rural residents’ basic medical insurance scheme (URRBMI) [1-2].

Voluntary enrollment is a principle of URRBMI in China [3]. The enrollee’s satisfaction and loyalty are crucial for the scheme’s effective implementation and sustained development. Moreover, analyzing the enrollee’s satisfaction with their health insurance and its influencing factors can help to shed light on current and future policy decision making [0, 4-4]. ACSI is one of the most widely used and representative quantitative measurement scales that shows the cause-and-effect relationship linking the casual variables of customer satisfaction to their consequent effect variable (customer loyalty) [6]. The model construction theory of ACSI provides an important reference for assessing the attitude and major improvement direction of the URRBMI from the perspective of insured participants. Using the ACSI as a basis and considering the Chinese social and cultural backgrounds, Peng et al. constructed a satisfaction index model for URRBMI that demonstrated good reliability and validity for satisfaction measurement[7-8]. There has been sufficient literature supporting the notion that perceived quality may be the most worthwhile topic in service area satisfaction research, and satisfaction is mainly determined by perceived quality [6, 9-10]. However, Peng’s model only adopted a few indicators to assess perceived quality, such as the overall quality assessment question, “how much help does the URRBMI bring to your child?”. The lack of comprehensive and detailed aspects of URRBMI quality measurement greatly limit its further use in health reform [7-8].

In addition, since significant variation in economic and cultural backgrounds, accessibility of health services, and reimbursement policies existed between rural and urban residents before the URRBMI integration, it is necessary to consider the modeling heterogeneity from a theoretical and practical perspective to learn about the differences between groups of the insured (rural VS urban) [11].

The aims of this study are to revise and validate the SIM-URRBMI and identify the main quality aspects needed to be improved in order to provide a valid, reliable, and practical satisfaction measurement tool for URRBMI. In addition, using heterogeneity analysis, we aim to explore the differences in satisfaction mechanism between rural and urban participants.

## Materials and Methods

### Participants and Sampling

This study was conducted in Changsha, the capital city of the Chinese Province of Hunan, which completed the integration of NCMS and URBMI in 2011. A three-stage randomized stratified cluster sampling method was used to ensure a representative sample. The first stage was to randomly select two districts/counties from the nine districts/counties of Changsha City. The second stage was to randomly select two primary schools for each selected district/county. Finally, for each selected school, six classes (one class from each grade) were randomly selected, and questionnaires were sent home; the main family decision maker responsible for the pupils’ URRBMI was instructed to fill out the questionnaires. The inclusion criteria were: a) had insurance for their children in 2017-2018; b) were inhabitants of a city or village in Changsha; c) were the main decision maker for pupils’ URRBMI in their family. Both the students in the selected classes and the main family decision makers responsible for the pupils’ URRBMI were subjects of investigation.

This study used a graded response model (GRM) to identify the measurement variables. In order to make the estimated parameters more accurate, a sample size of at least 500 cases is suitable. The selected samples of measurement variables included in this paper are 574 cases, which meet the requirements[13]. The sample size for model evaluation was calculated by the following formula, *n* ≥ 50*r*^2^-450*r* + 1100 (*r* is the ratio of measurement variables to latent variables), which was based on the structural equation model research conducted by Westland et al. A minimum sample size of 216 was estimated. However, considering the need for subgroup analysis, a sample size of 1335 was obtained for model evaluation, which met the estimated requirements [13-14].

### Ethical approval

The study protocol was reviewed and approved by the Medical Ethics Committee at Xiang -ya School of Public Health, Central South University. Written informed consent was obtai ned for each participant who was the medical insurance decision maker of the family.

### Model construction

#### Phase 1

The programmed decision process, a method to simultaneously develop a scale by nominal group (including pupils and their family members, local medical insurance management and service officers, experts of social medicine, health management, primary health care, health statistics, and epidemiology et al.) and focus group (1 project leader, 1 professor of epidemiology and health statistics, 1 health insurance administration, and graduate students), was used to construct the model. With reference to the literature, we refined the “perceived quality” variable to five first-order latent variables, namely, overall quality, information quality, service quality, policy quality, and institution quality. The draft model consisted of 11 latent variables, including public expectations (PE), perceived quality (PQ) (which was a second-order latent variable and consisted of the five first-order latent variables), perceived value (PV), public satisfaction (PS), public complaints (PC), and public trust (PT). Public expectation, perceived quality, and perceived value were the explanatory variables of public satisfaction, while public complaints and public trust were the outcome variables of public satisfaction. Consistent with Peng’s research, we used the term “public” as an alternative of the term “customer” and changed “customer loyalty” to “public trust”. Then, an initial draft variable pool (29 variables) was generated (see Table 1). Since the path of public complaints on public trust was not significant according to Peng’s research, this path was not set in the initial draft SIM-URRBMI.

**Table 1.**
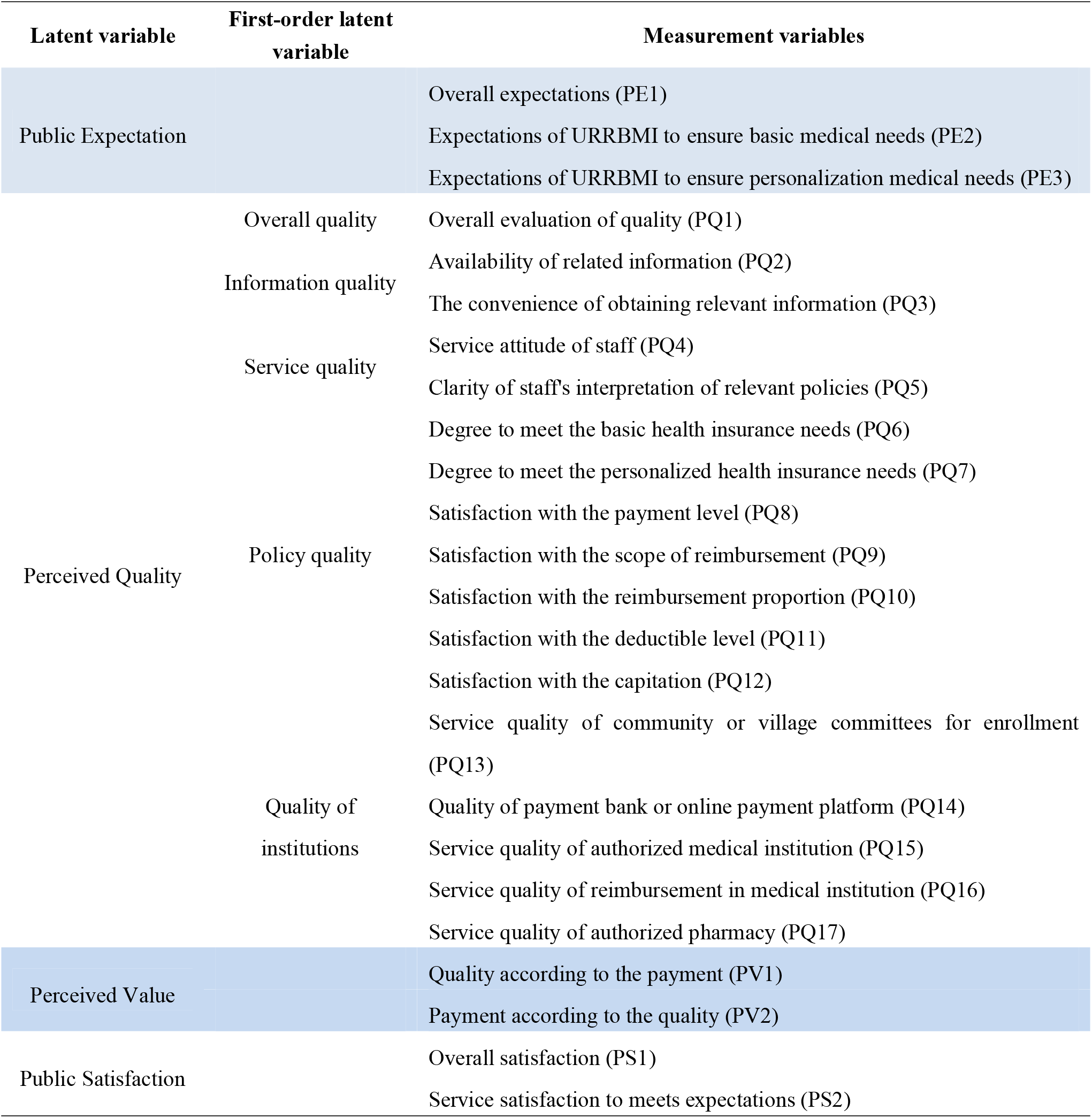
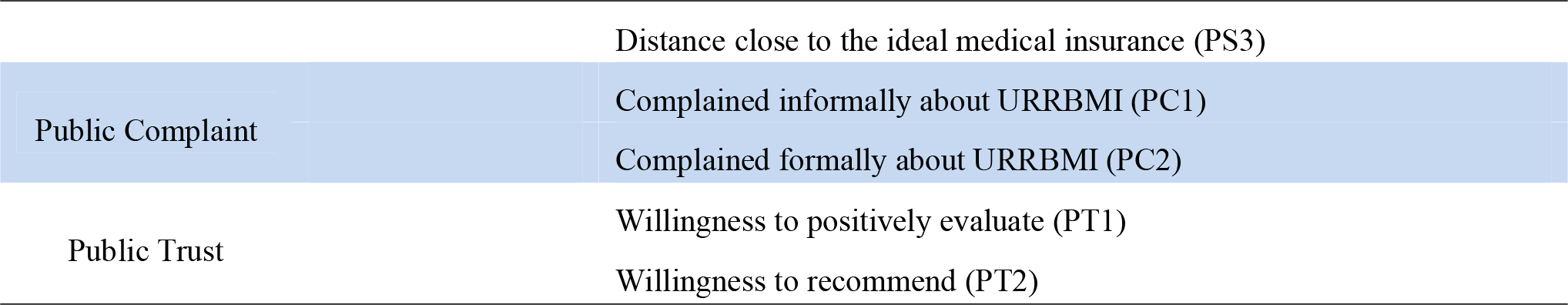
The latent and measurement variables of the initial SIM_URRBMI

Based on expert consultation and literature review, the following path hypotheses were established in the study (Fig 1):

Hypothesis 1: Public expectation has a direct positive impact on perceived quality, perceived value, and public satisfaction;
Hypothesis 2: Perceived quality has a direct positive impact on perceived value and public satisfaction;
Hypothesis 3: Perceived value has a direct positive impact on public satisfaction;
Hypothesis 4: Public satisfaction has a direct positive impact on public trust;
Hypothesis 5: Public satisfaction has a direct negative impact on public complaints.

**Fig1.**
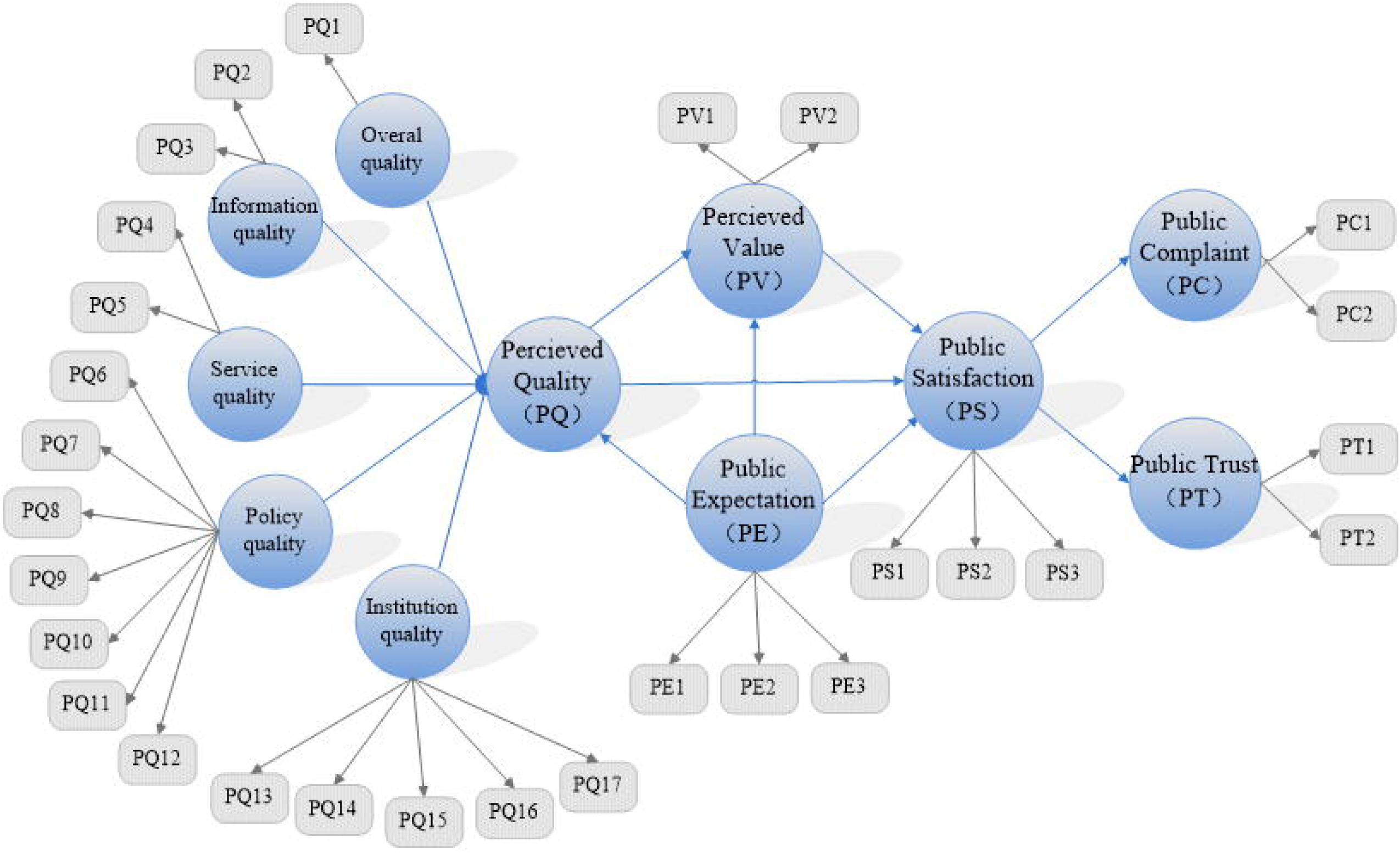
The initial draft of SIM_URRBMI

#### Phase 2

Classic Test Theory (CTT) combined with Item Response Theory (IRT) was applied for the measurement variable selection. If a measurement variable reached 3 or more deletion criteria, it was considered for deletion. CTT is based on the true score model (true score plus error), and its selection methods and criteria are shown in Table 2. The basic idea of IRT is to use a mathematical function to characterize the relationship between the test response; in this study, *θ* refers to subject satisfaction with URRBMI. For the graded response model (GRM) in this study, the discrimination parameter (*a*), Threshold Parameter (*b*), and Item Information Function (*IIF*) were used to evaluate the quality of measurement variables. In addition, the Test Information Function (*TIF*) is a linear cumulative measurement of each item *IIF*; that is, the test information function on the *θ* value is equal to the sum of its *IIFs*. It is generally considered that the quality of the model is good when the total of the information function is greater than 25, while the quality is considered poor when the information function is less than 16 [15-16]. Since the initial draft of the SIM-URRBMI contains a total of 29 measurement variables, the average information function amount of each measurement variable required 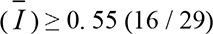. The deletion criteria based on IRT are as follows: (1) *a* is less than 0.3; (2) *b* is out of range of (−4,4); and (3) 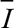 is less than 0. 55.

**Table 2.**
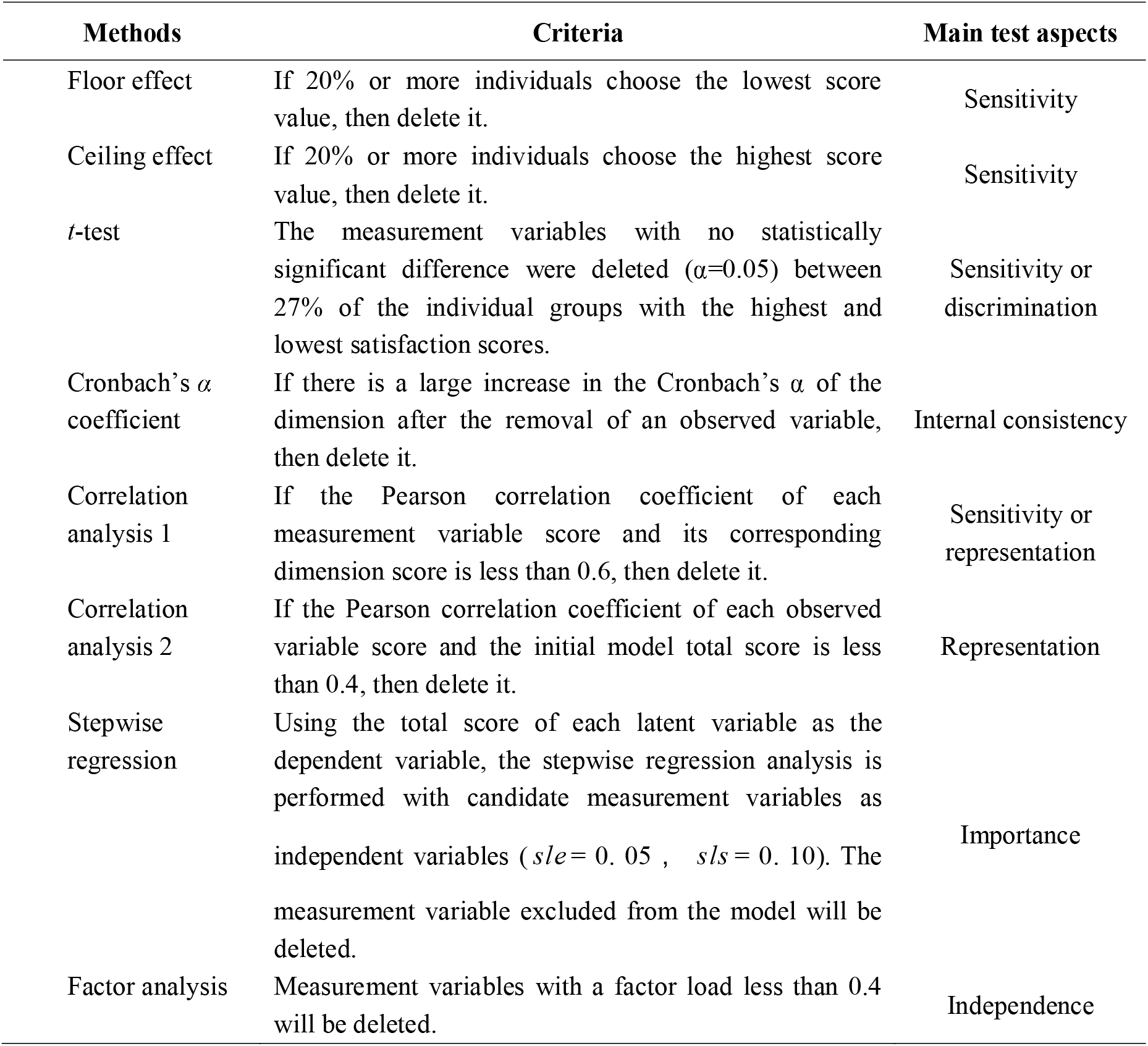
Item selection methods and criteria based on CTT

### Model validation

The model validation includes two parts. The major reliability and validity tests were used to evaluate the measurement model. The value of Cronbach’s α coefficient and the composite reliability coefficient were greater than 0.7. It is considered to indicate good reliability, and higher values indicate greater homogeneity. Outer loading (≥0.708) and Average Variance Extraction (*AVE*) (≥0.5) were used to assess the convergence validity, while the Cross Loading and Fornell-Larcker criteria were used to assess discriminant validity. Good discriminant validity means that the outer loading of a measurement variable on its corresponding latent variable is greater than its cross loadings on other latent variables, or the square root of the *AVE* of each latent variable is greater than its linear correlation coefficients with any other latent variables. Path analysis of the latent variables and model prediction were used to evaluate the structural model [11]. The relationship between latent variables was evaluated by estimating direct effect, indirect effect, and total effect. The model prediction assessment indicators were the adjusted coefficient of determination 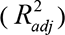 (In general, 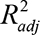values of 0.25, 0.50, and 0.75 for target constructs are considered weak, medium, and substantial, respectively.), the effect size (*f*^2^) (Results of 0.02, 0.15, and 0.35 are interpreted as small, medium, and large *f*^2^ effect sizes, respectively.), and the predictive relevance (*Q*^2^) (The path model has predictive relevance for a selected endogenous construct if the *Q*^2^ value is above zero).

### Statistical analysis

Second-order Partial Least Squares Structural Equation Model (PLS-SEM) was used to test the model’s reliability and validity, and PLS-SEM Multigroup Analysis (PLS-MGA) was used to explore the heterogeneity between rural and urban participants. A two-tailed *P*≤ 0.05 was considered statistically significant. To be consistent with the international customer satisfaction index, a one hundred percentage point system was used to express the public satisfaction index (*PSI*).

The descriptive statistics and CTT analyses were conducted using SPSS 18.0. IRT analysis was performed using Mplus version 7.0. PLS-SEM and PLS-MGA analyses were conducted with SmartPLS 3.0.

## Results

### Samples

A total of 1909 participants who had insurance for their children were included in the primary data analysis. Table 3 shows the sociodemographic information of pupils and main decision makers for the model construction group and model validation group separately. For pupils, the mean age was 9.26±1.75, and the frequency of male (945, 49.89%) and female students (949, 50.11%) were similar. For the main decision makers, the mean age was 37.13±5.90 years; most of decision makers were parents of pupils (1845, 96.95%), especially mothers (1196, 62.85%). Table 3 also indicates that there were no significant differences between the two groups for both pupils and main decision makers, suggesting that the two groups were homogenous. The measurement scores and comparative analysis between the two groups are shown in S1 Table.

**Table 3.**
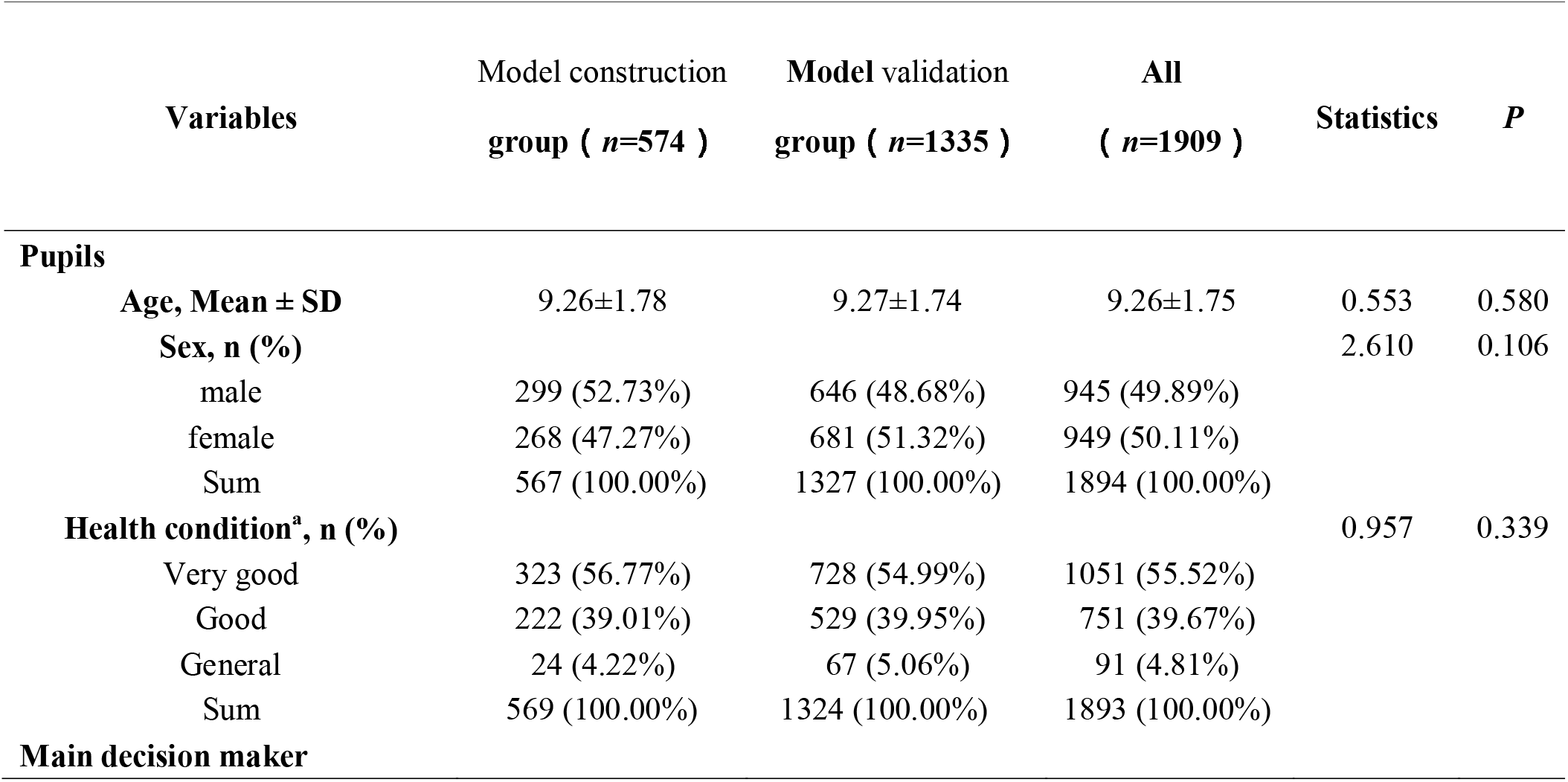
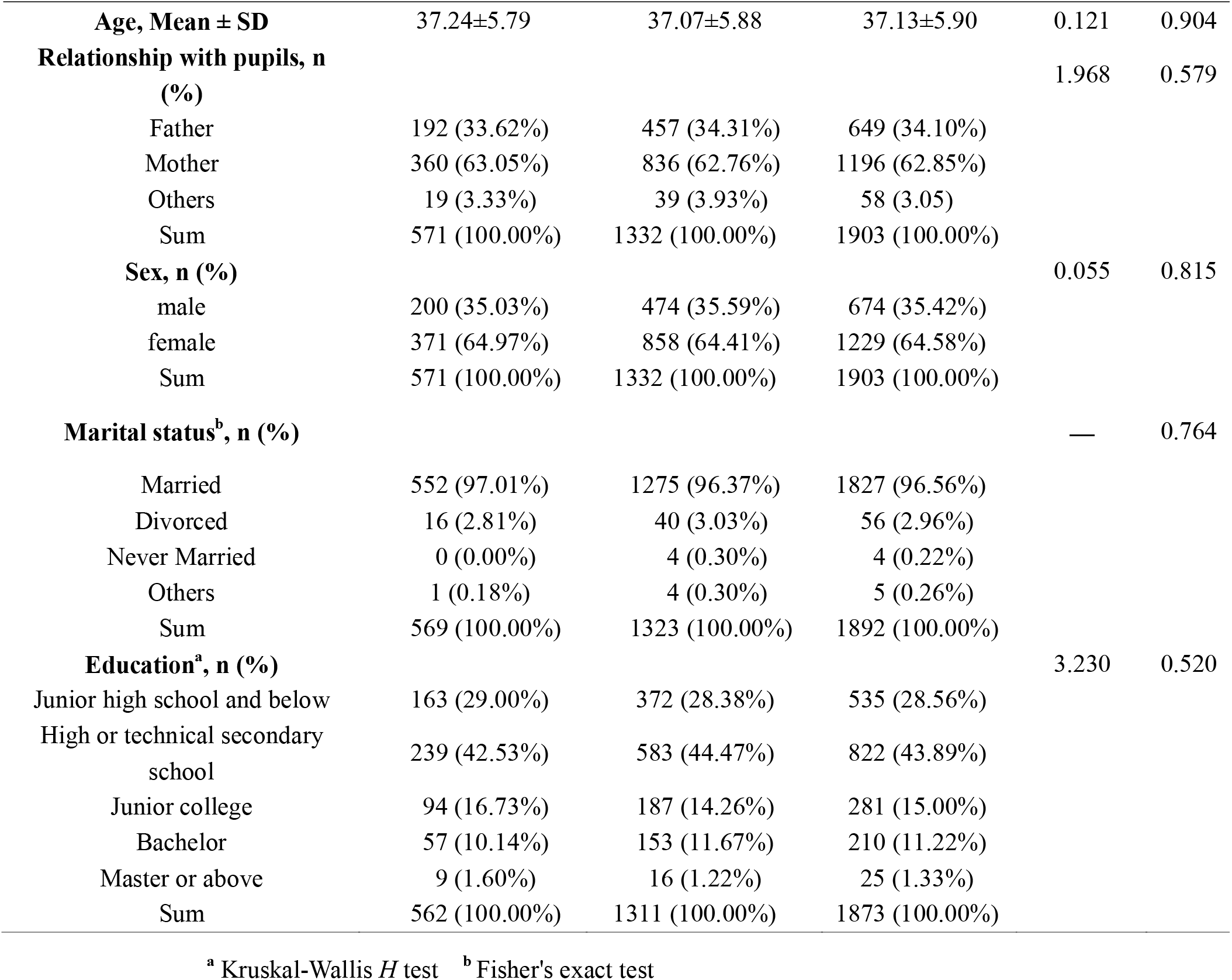
Sociodemographic information of the pupils and the main decision makers for their BMI

| Variables | Model construction<br>group ( *n*=574 ) | Model validation<br>group ( *n*=1335 ) | All<br>( *n*=1909 ) | Statistics | *P* |
| --- | --- | --- | --- | --- | --- |
| **Pupils** |  |  |  |  |  |
| Age, Mean ± SD | 9.26±1.78 | 9.27±1.74 | 9.26±1.75 | 0.553 | 0.580 |
| Sex, *n* (%) |  |  |  | 2.610 | 0.106 |
| male | 299 (52.73%) | 646 (48.68%) | 945 (49.89%) |  |  |
| female | 268 (47.27%) | 681 (51.32%) | 949 (50.11%) |  |  |
| Sum | 567 (100.00%) | 1327 (100.00%) | 1894 (100.00%) |  |  |
| Health condition<sup>a</sup>, *n* (%) |  |  |  | 0.957 | 0.339 |
| Very good | 323 (56.77%) | 728 (54.99%) | 1051 (55.52%) |  |  |
| Good | 222 (39.01%) | 529 (39.95%) | 751 (39.67%) |  |  |
| General | 24 (4.22%) | 67 (5.06%) | 91 (4.81%) |  |  |
| Sum | 569 (100.00%) | 1324 (100.00%) | 1893 (100.00%) |  |  |
| **Main decision maker** |  |  |  |  |  |
| **Age, Mean ± SD** | 37.24±5.79 | 37.07±5.88 | 37.13±5.90 | 0.121 | 0.904 |
| **Relationship with pupils, n (%)** |  |  |  | 1.968 | 0.579 |
| Father | 192 (33.62%) | 457 (34.31%) | 649 (34.10%) |  |  |
| Mother | 360 (63.05%) | 836 (62.76%) | 1196 (62.85%) |  |  |
| Others | 19 (3.33%) | 39 (3.93%) | 58 (3.05%) |  |  |
| Sum | 571 (100.00%) | 1332 (100.00%) | 1903 (100.00%) |  |  |
| **Sex, n (%)** |  |  |  | 0.055 | 0.815 |
| male | 200 (35.03%) | 474 (35.59%) | 674 (35.42%) |  |  |
| female | 371 (64.97%) | 858 (64.41%) | 1229 (64.58%) |  |  |
| Sum | 571 (100.00%) | 1332 (100.00%) | 1903 (100.00%) |  |  |
| **Marital status<sup>b</sup>, n (%)** |  |  |  | — | 0.764 |
| Married | 552 (97.01%) | 1275 (96.37%) | 1827 (96.56%) |  |  |
| Divorced | 16 (2.81%) | 40 (3.03%) | 56 (2.96%) |  |  |
| Never Married | 0 (0.00%) | 4 (0.30%) | 4 (0.22%) |  |  |
| Others | 1 (0.18%) | 4 (0.30%) | 5 (0.26%) |  |  |
| Sum | 569 (100.00%) | 1323 (100.00%) | 1892 (100.00%) |  |  |
| **Education<sup>a</sup>, n (%)** |  |  |  | 3.230 | 0.520 |
| Junior high school and below | 163 (29.00%) | 372 (28.38%) | 535 (28.56%) |  |  |
| High or technical secondary school | 239 (42.53%) | 583 (44.47%) | 822 (43.89%) |  |  |
| Junior college | 94 (16.73%) | 187 (14.26%) | 281 (15.00%) |  |  |
| Bachelor | 57 (10.14%) | 153 (11.67%) | 210 (11.22%) |  |  |
| Master or above | 9 (1.60%) | 16 (1.22%) | 25 (1.33%) |  |  |
| Sum | 562 (100.00%) | 1311 (100.00%) | 1873 (100.00%) |  |  |

<sup>a</sup> Kruskal-Wallis $H$ test <sup>b</sup> Fisher's exact test

### Model construction

S2 Table provided GRM estimations of the discrimination (a) and threshold parameters (*b*1-*b*9) as well as their standard errors (SE). Table 4 shows the results of measurement variable selection with the CTT and IRT methods. Using these methods, the ceiling effect selected 5 variables (PQ1, PQ14, PE2, PT1 and PT2), Cronbach’s *α* coefficient selected 1 variable (PS3), a selected 1 variable (PC1), *b* selected 9 variables (PQ9-PQ12, PQ16, PV1, PV2, PS1 and PS2), and 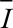 selected 4 variables (PQ1, PQ2, PC1, and PC2).

**Table 4.**
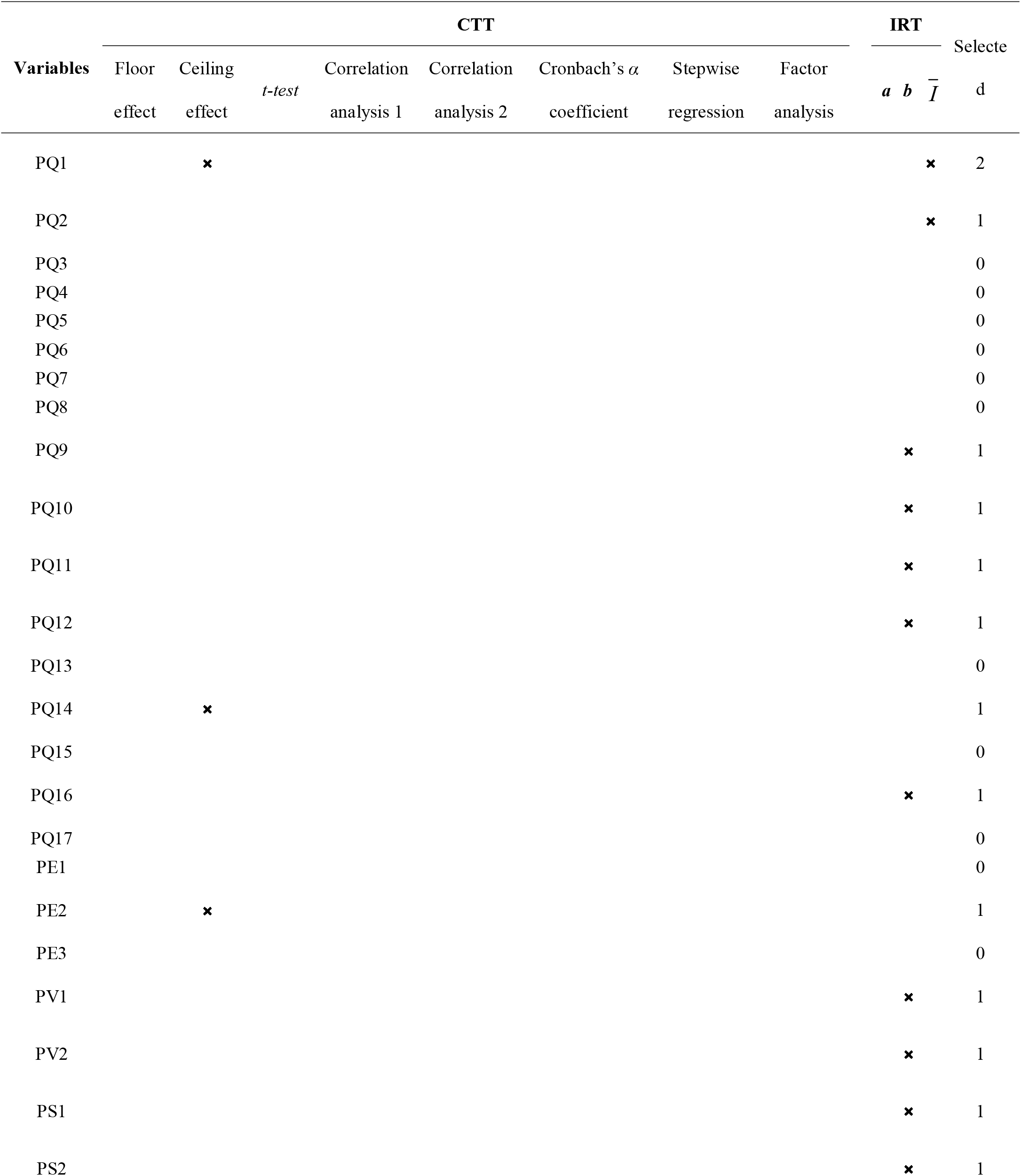
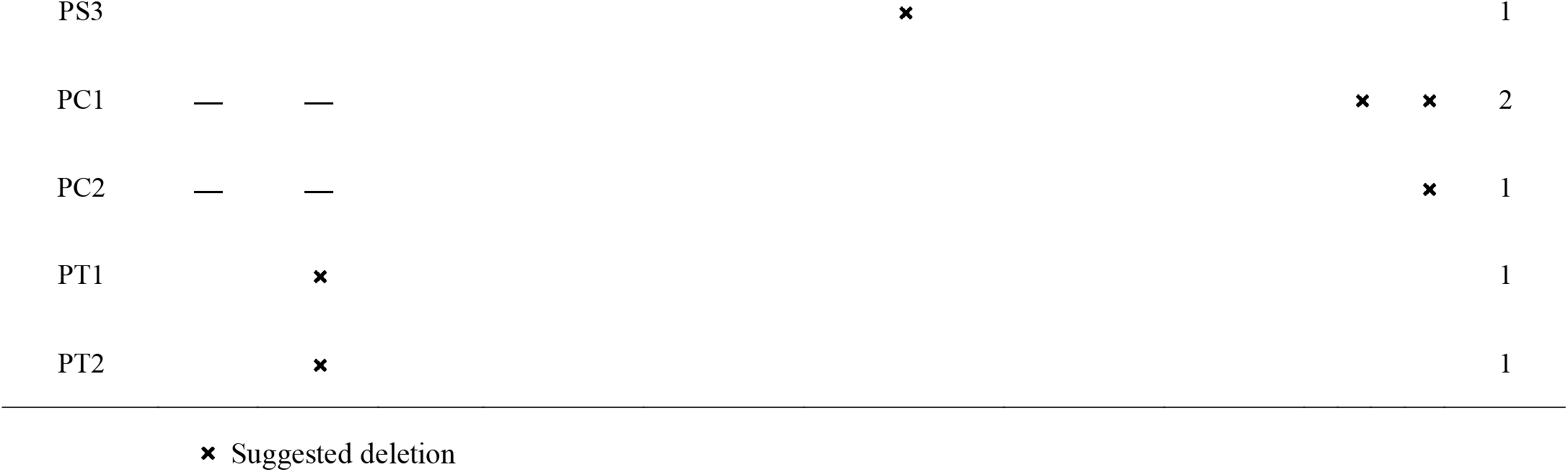
The results of measurement variable selection with the methods of CTT and IRT

Overall, no item reached deletion criteria 3 or more times. However, it can be seen that the 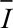 of PC2 is 0.005, suggesting that its contribution to the model is near 0, and its discrimination is also low (*a* is 0.351). In addition, with China’s social and cultural backgrounds, most enrollees will not formally complain even if they are dissatisfied. Thus, the PC2 was deleted, and the remaining 28 variables were used to construct the SIM-URRBMI (Fig 2).

**Fig2.**
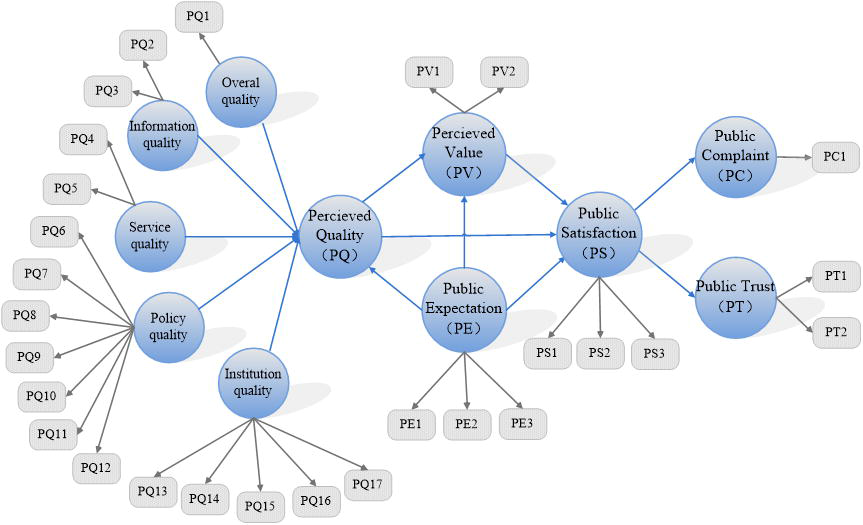
The final version of SIM_URRBMI

### Model validation

#### Reliability

The Cronbach’s *α* coefficient for each of the latent variables ranged from 0.772 to 1.000, and the composite reliability coefficients ranged from 0.897 to 1.000, both of which are greater than 0.7, providing evidence that the revised SIM-URRBMI is reliable (see Table 5).

**Table 5.**
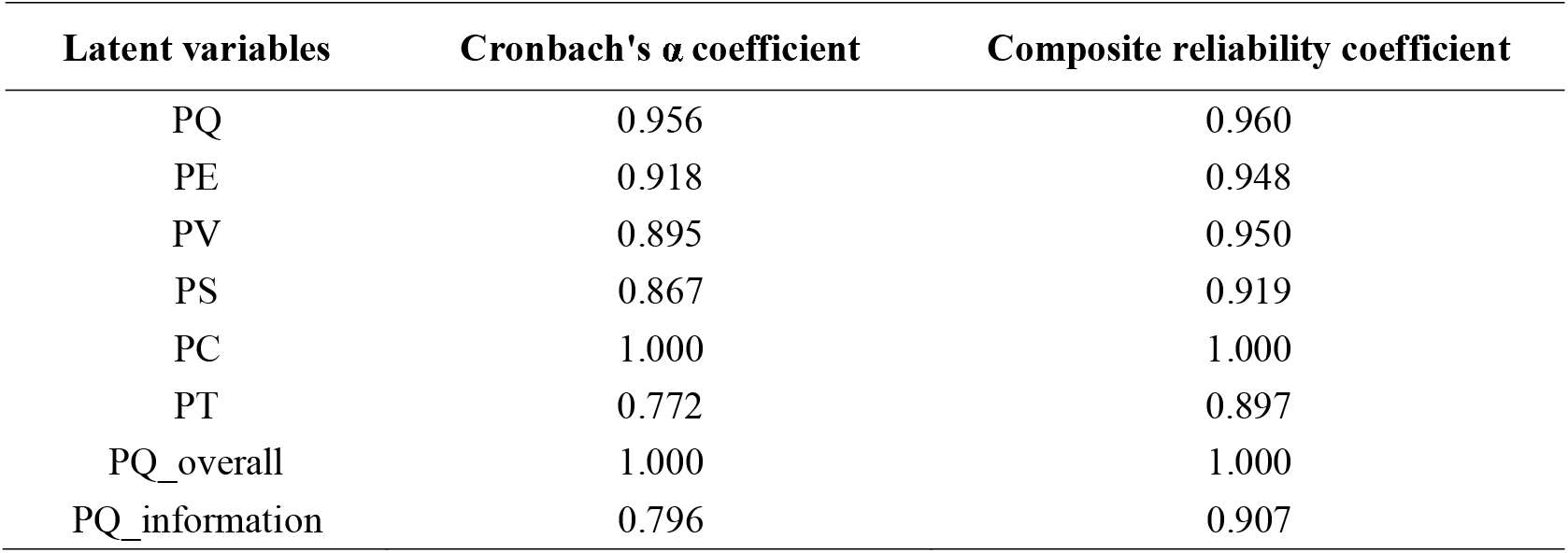
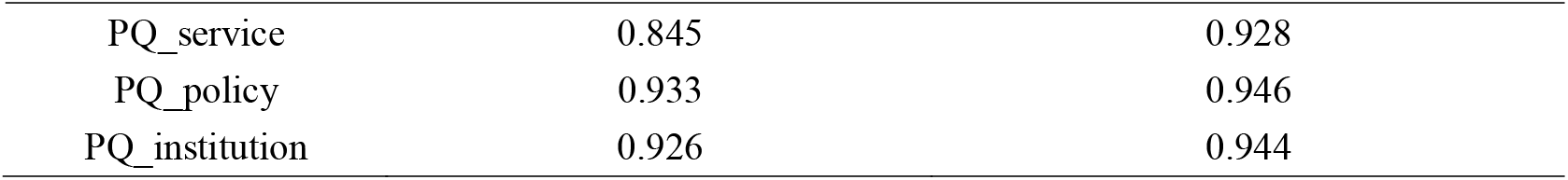
The reliability of the SIM-URRBMI

#### Validity

The results of the convergent and discriminant validity of the revised SIM-URRBMI are summarized in Tables 6 - 8. The *AVEs* of all latent variables were greater than 0.5, and the outer factor loadings of all the measurement variables ranged from 0.726 to 1.000, except that the second-order external factor loadings for the PQ1, PQ2, and PQ3 variables (0.692, 0.595 and 0.684, respectively) was slightly below the critical value of 0.7, showing good convergent validity. All of the outer loadings of the measurement variables on their corresponding latent variable were greater than their cross loadings on the other latent variables. The square roots of the *AVEs* for each latent variable were greater than the linear correlation of the latent variable with the other structures, except for the perceived quality variable, which has a slightly higher correlation with perceived value and public satisfaction(Table 8). The results indicate good discriminant validity. All but one of the paths were statistically significant and positive; the effect of public satisfaction on the public complaints was negative, which is in line with the model theory framework. Table 9 and S3 Table show the result of the direct effects and the collinearity assessment of the SIM-URRBMI. Furthermore, the greatest effect of the five first-order latent variables on perceived quality was medical insurance policy quality (0.472) followed by institution quality (0.354) (see Table 9). All variance inflation factor (*VIF*) values are clearly below the threshold of 5. Therefore, collinearity among the predictor constructs is not an issue in the structural model. The two paths with the greatest direct effect were perceived quality on perceived value (0.676) and public satisfaction on public trust (0.634). S4 Table indicates the indirect effects of the revised SIM-URRBMI.

**Table 6.**
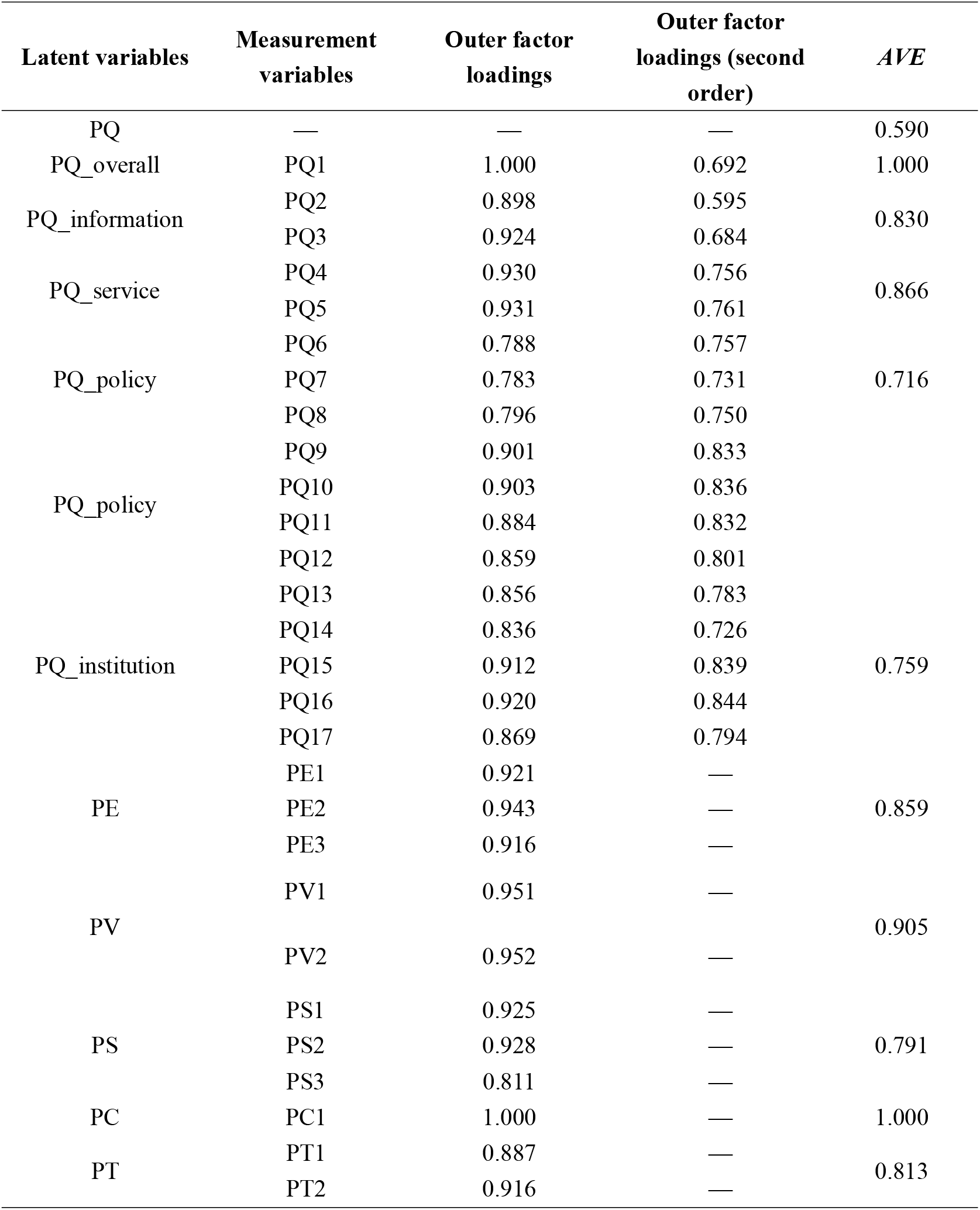
The convergent validity of the SIM-URRBMI

**Table 7.** Outer factor loadings and cross loadings of measurement variables for the SIM-URRBMI

| Variables | PQ_<br>overall<br>1 | PQ_<br>information | PQ_<br>service | PQ_<br>policy | PQ_<br>institution | PQ<br>(second<br>order) | PE | PV | PS | PC | PT |
| --- | --- | --- | --- | --- | --- | --- | --- | --- | --- | --- | --- |
| PQ1 | **1.000** | 0.499 | 0.582 | 0.586 | 0.597 | **0.692** | 0.427 | 0.561 | 0.584 | -0.224 | 0.474 |
| PQ2 | 0.428 | **0.898** | 0.496 | 0.515 | 0.439 | **0.595** | 0.287 | 0.444 | 0.460 | -0.118 | 0.305 |
| PQ3 | 0.479 | **0.924** | 0.619 | 0.588 | 0.529 | **0.684** | 0.333 | 0.482 | 0.515 | -0.132 | 0.375 |
| PQ4 | 0.577 | 0.544 | **0.930** | 0.595 | 0.703 | **0.756** | 0.440 | 0.565 | 0.597 | -0.216 | 0.467 |
| PQ5 | 0.507 | 0.603 | **0.931** | 0.645 | 0.645 | **0.761** | 0.434 | 0.568 | 0.604 | -0.204 | 0.416 |
| PQ6 | 0.533 | 0.546 | 0.610 | **0.788** | 0.584 | **0.757** | 0.448 | 0.596 | 0.624 | -0.196 | 0.439 |
| PQ7 | 0.442 | 0.537 | 0.566 | **0.783** | 0.556 | **0.731** | 0.389 | 0.540 | 0.591 | -0.190 | 0.392 |
| PQ8 | 0.505 | 0.478 | 0.514 | **0.796** | 0.612 | **0.750** | 0.455 | 0.671 | 0.633 | -0.267 | 0.524 |
| PQ9 | 0.521 | 0.516 | 0.578 | **0.901** | 0.666 | **0.833** | 0.394 | 0.674 | 0.698 | -0.260 | 0.476 |
| PQ10 | 0.512 | 0.500 | 0.565 | **0.903** | 0.681 | **0.836** | 0.421 | 0.680 | 0.708 | -0.238 | 0.520 |
| PQ11 | 0.494 | 0.522 | 0.570 | **0.884** | 0.690 | **0.832** | 0.409 | 0.681 | 0.684 | -0.262 | 0.504 |
| PQ12 | 0.463 | 0.502 | 0.544 | **0.859** | 0.658 | **0.801** | 0.366 | 0.652 | 0.672 | -0.280 | 0.481 |
| PQ13 | 0.490 | 0.456 | 0.675 | 0.644 | **0.856** | **0.783** | 0.493 | 0.590 | 0.650 | -0.213 | 0.517 |
| PQ14 | 0.507 | 0.418 | 0.583 | 0.573 | **0.836** | **0.726** | 0.478 | 0.565 | 0.597 | -0.193 | 0.503 |
| PQ15 | 0.546 | 0.505 | 0.653 | 0.706 | **0.912** | **0.839** | 0.521 | 0.678 | 0.735 | -0.237 | 0.568 |
| PQ16 | 0.553 | 0.507 | 0.663 | 0.706 | **0.920** | **0.844** | 0.534 | 0.693 | 0.747 | -0.255 | 0.575 |
| PQ17 | 0.526 | 0.458 | 0.608 | 0.671 | **0.869** | **0.794** | 0.535 | 0.629 | 0.671 | -0.207 | 0.539 |
| PE1 | 0.397 | 0.332 | 0.449 | 0.448 | 0.559 | 0.534 | **0.921** | 0.537 | 0.546 | -0.083 | 0.486 |
| PE2 | 0.391 | 0.297 | 0.444 | 0.437 | 0.542 | 0.517 | **0.943** | 0.547 | 0.541 | -0.074 | 0.491 |
| PE3 | 0.400 | 0.322 | 0.411 | 0.468 | 0.520 | 0.524 | **0.916** | 0.544 | 0.531 | -0.052 | 0.454 |
| PV1 | 0.540 | 0.484 | 0.577 | 0.726 | 0.685 | 0.755 | 0.556 | **0.951** | 0.795 | -0.293 | 0.601 |
| PV2 | 0.528 | 0.485 | 0.581 | 0.721 | 0.684 | 0.752 | 0.557 | **0.952** | 0.807 | -0.264 | 0.589 |
| PS1 | 0.581 | 0.493 | 0.629 | 0.741 | 0.754 | 0.797 | 0.559 | 0.831 | **0.925** | -0.297 | 0.598 |
| PS2 | 0.544 | 0.513 | 0.616 | 0.743 | 0.729 | 0.787 | 0.528 | 0.791 | **0.928** | -0.276 | 0.563 |
| PS3 | 0.419 | 0.420 | 0.462 | 0.583 | 0.571 | 0.615 | 0.461 | 0.604 | **0.811** | -0.212 | 0.526 |
| PC1 | -0.275 | -0.169 | -0.277 | -0.351 | -0.310 | -0.351 | -0.093 | -0.359 | -0.364 | **1.000** | -0.251 |
| PT1 | 0.389 | 0.298 | 0.390 | 0.472 | 0.520 | 0.520 | 0.456 | 0.509 | 0.530 | -0.142 | **0.887** |
| PT2 | 0.462 | 0.376 | 0.461 | 0.542 | 0.587 | 0.600 | 0.472 | 0.613 | 0.607 | -0.223 | **0.916** |

The bold number in the table is the external factor loadings of the measurement variable on its
corresponding latent variable, and the rest are the cross loadings.

**Table 8.** The Fornell-Larcker standard for the SIM-URRBMI

|  | PQ | PQ_<br>overall<br>1 | PQ_<br>information | PQ_<br>service | PQ_<br>policy | PQ_<br>institution | PE | PV | PS | PC | PT |
| --- | --- | --- | --- | --- | --- | --- | --- | --- | --- | --- | --- |
| PQ | **0.768** |  |  |  |  |  |  |  |  |  |  |
| PQ_<br>overall | 0.692 | **1.000** |  |  |  |  |  |  |  |  |  |
| PQ_<br>Information | 0.705 | 0.499 | **0.911** |  |  |  |  |  |  |  |  |
| PQ_<br>service | 0.815 | 0.582 | 0.616 | **0.931** |  |  |  |  |  |  |  |
| PQ_<br>policy | 0.937 | 0.586 | 0.608 | 0.666 | **0.846** |  |  |  |  |  |  |
| PQ_<br>institution | 0.908 | 0.597 | 0.534 | 0.724 | 0.753 | **0.879** |  |  |  |  |  |
| PE | 0.567 | 0.427 | 0.342 | 0.470 | 0.486 | 0.583 | **0.927** |  |  |  |  |
| PV | 0.792 | 0.561 | 0.510 | 0.609 | 0.760 | 0.720 | 0.585 | **0.951** |  |  |  |
| PS | 0.830 | 0.584 | 0.536 | 0.646 | 0.780 | 0.776 | 0.582 | 0.842 | **0.890** |  |  |
| PC | -0.286 | -0.224 | -0.138 | -0.226 | -0.286 | -0.252 | -0.076 | -0.293 | -0.297 | **1.000** |  |
| PT | 0.623 | 0.474 | 0.376 | 0.475 | 0.565 | 0.616 | 0.515 | 0.625 | 0.633 | -0.205 | **0.902** |

The bolded numbers in the diagonal of the table are the square roots of *AVE* for each latent variable

**Table 9.** The direct effects in the SIM-URRBMI

| Direct path | Coefficient | SE | *t* | *P* | 95%CI |  |
| --- | --- | --- | --- | --- | --- | --- |
|  |  |  |  |  | Lower | Upper |
| PE→PQ | 0.568 | — | — | — | — | — |
| PE→PV | 0.204 | 0.027 | 7.372 | <0.001 | 0.145 | 0.253 |
| PE→PS | 0.070 | 0.021 | 3.362 | 0.001 | 0.034 | 0.119 |
| PQ→PV | 0.676 | 0.021 | 31.964 | <0.001 | 0.639 | 0.720 |
| PQ→PS | 0.421 | 0.028 | 14.960 | <0.001 | 0.361 | 0.474 |
| PV→PS | 0.467 | 0.032 | 14.698 | <0.001 | 0.404 | 0.528 |
| PS→PC | -0.243 | 0.020 | 11.847 | <0.001 | -0.280 | -0.202 |
| PS→PT | 0.634 | 0.020 | 31.305 | <0.001 | 0.596 | 0.674 |
| PE→PQ_overall | 0.428 | 0.027 | 15.651 | <0.001 | 0.373 | 0.477 |
| PE→PQ_information | 0.344 | 0.026 | 12.981 | <0.001 | 0.290 | 0.393 |
| PE→PQ_service | 0.471 | 0.026 | 18.247 | <0.001 | 0.413 | 0.518 |
| PE→PQ_policy | 0.487 | 0.025 | 19.415 | <0.001 | 0.437 | 0.534 |
| PE→PQ_institution | 0.584 | 0.023 | 25.431 | <0.001 | 0.534 | 0.624 |
| PQ_overall→PQ | 0.069 | 0.002 | 36.692 | <0.001 | 0.066 | 0.073 |
| PQ_information→PQ | 0.110 | 0.003 | 31.786 | <0.001 | 0.103 | 0.117 |
| PQ_service→PQ | 0.135 | 0.003 | 47.898 | <0.001 | 0.130 | 0.142 |
| PQ_policy→PQ | 0.472 | 0.006 | 76.267 | <0.001 | 0.462 | 0.485 |
| PQ_institution→PQ | 0.354 | 0.005 | 64.894 | <0.001 | 0.343 | 0.364 |

The total effect of the 14 paths of the SIM-URRBMI were statistically significant, showing that the model was constructed properly (see Table 10). The three strongest paths were (1) perceived quality on public satisfaction (with a total effect of 0.737, a direct effect of 0.421 and an indirect effect of 0.316); (2) perceived quality on perceived value (with a total effect of 0.676);and (3) public satisfaction on public trust (with a total effect of 0.634). The largest total effect of the five explanatory variables of public trust was calculated for public satisfaction (0.634), followed by perceived quality (0.467). Among the three explanatory variables of public satisfaction, the greatest total effect was from perceived quality (0.737).

**Table 10.** The results of the total effect analysis for the SIM-URRBMI

| Total path | Coefficient | SE | t | P | 95%CI |  |
| --- | --- | --- | --- | --- | --- | --- |
|  |  |  |  |  | Lower | Upper |
| PE→PQ<sup>a</sup> | 0.568 | 0.022 | 25.377 | <0.001 | 0.519 | 0.609 |
| PE→PQ | 0.588 | 0.025 | 23.122 | <0.001 | 0.535 | 0.637 |
| PE→PS | 0.584 | 0.024 | 23.953 | <0.001 | 0.535 | 0.630 |
| PE→PC<sup>b</sup> | -0.142 | 0.013 | 10.963 | <0.001 | -0.168 | -0.116 |
| PE→PT<sup>b</sup> | 0.371 | 0.023 | 16.142 | <0.001 | 0.322 | 0.409 |
| PQ→PV<sup>a</sup> | 0.676 | 0.021 | 31.964 | <0.001 | 0.639 | 0.720 |
| PQ→PS | 0.737 | 0.018 | 41.270 | <0.001 | 0.699 | 0.771 |
| PQ→PC<sup>b</sup> | -0.179 | 0.016 | 10.870 | <0.001 | -0.210 | -0.143 |
| PQ→PT<sup>b</sup> | 0.467 | 0.018 | 25.732 | <0.001 | 0.430 | 0.502 |
| PV→PS<sup>a</sup> | 0.467 | 0.032 | 14.698 | <0.001 | 0.404 | 0.528 |
| PV→PC<sup>b</sup> | -0.113 | 0.013 | 9.010 | <0.001 | -0.139 | -0.090 |
| PV→PT<sup>b</sup> | 0.297 | 0.023 | 13.161 | <0.001 | 0.252 | 0.336 |
| PS→PC<sup>a</sup> | -0.243 | 0.020 | 11.847 | <0.001 | -0.280 | -0.202 |
| PS→PT | 0.634 | 0.020 | 31.305 | <0.001 | 0.596 | 0.674 |

<sup>a</sup> indicates that only the direct path was set, <sup>b</sup> indicates that only the indirect path was set

The adjusted coefficients of determination 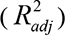 of each endogenous latent variable ranged from 0.088 to 1.000. The 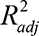 values were 0.783 and 0.400 for public satisfaction and public trust, respectively, indicating a strong and moderate prediction. The effect size (*f*^2^) ranged from 0.014 to 0.903; the *f*^2^ values for perceived quality on predicted perceived value, the perceived value on predicted public satisfaction, and public satisfaction on predicted public trust were 0.903, 0.351 and 0.667, respectively; all were greater than 0.35 indicating a strong predictive effect. The predictive relevance (*Q*^2^) of all endogenous latent variables was greater than 0, ranging from 0.105 to 0.585, showing a certain predictive relevance for all five endogenous latent variables (see Table 11).

**Table 11.**
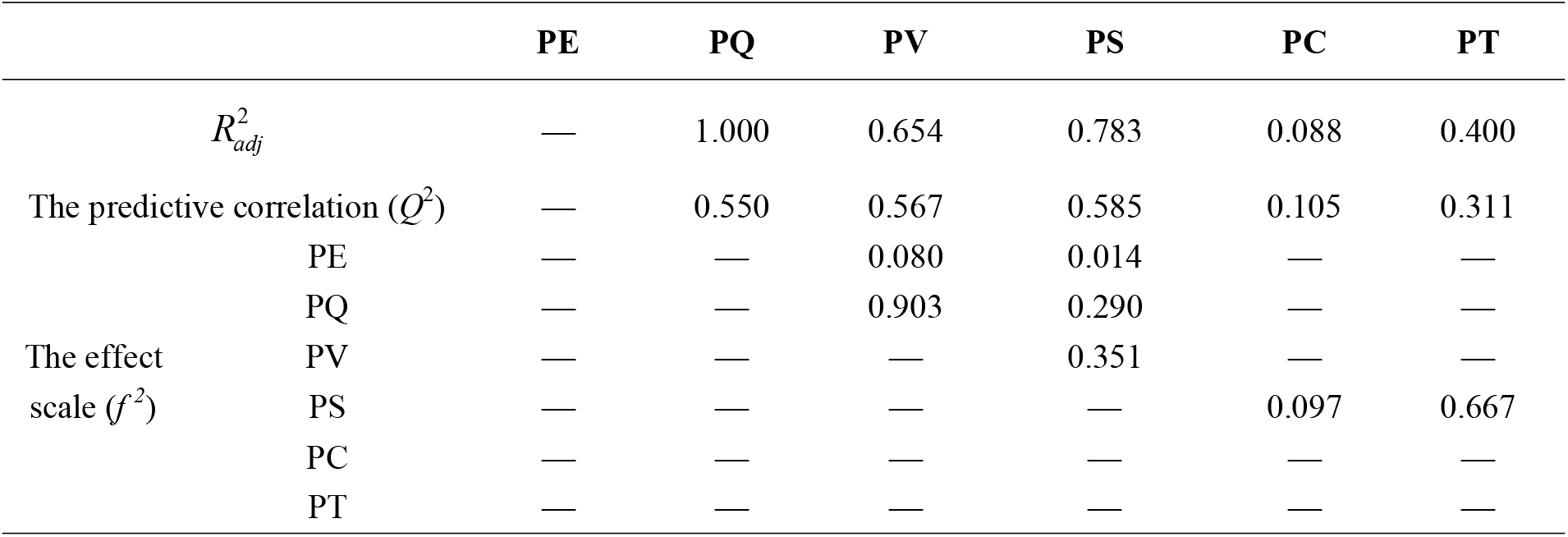
The results of predictive ability evaluation for the SIM-URRBMI

### Model application

The SIM-URRBMI satisfaction index scores were 57.29 and 58.17 for rural and urban participants, respectively. PLS-MGA showed that the total effect of public expectation on public satisfaction and public complaints was higher for citizens than for rural residents (0.627 vs 0.534, *P*=0.037; -0.166 vs -0.114, *P*=0.042). The total effect of perceived quality on public trust for citizens was lower than that of rural residents (0.429 vs 0.514, *P*=0.023) (see Table 12).

**Table12.**
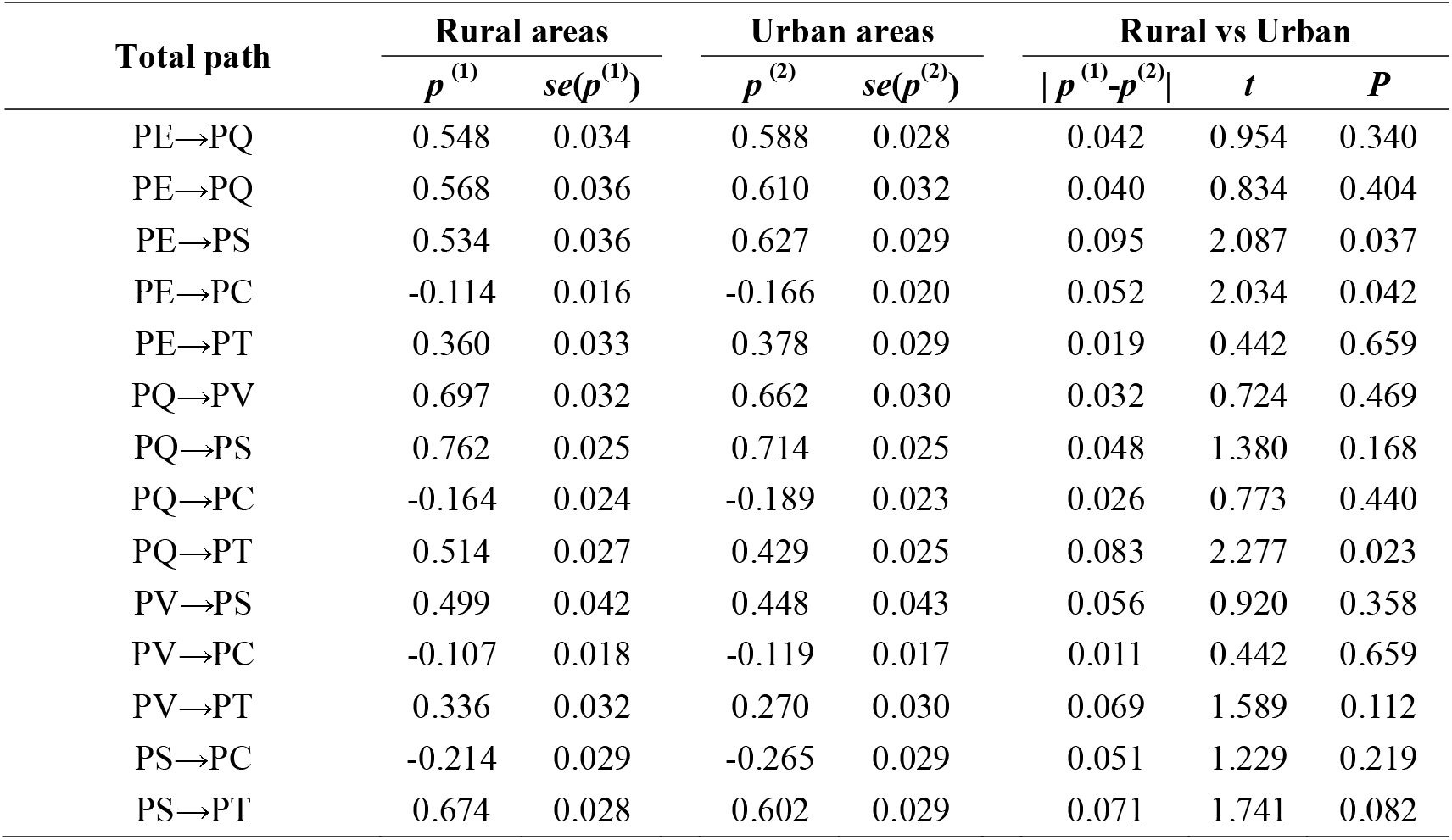
Comparison of total effects between urban and rural residents’ among latent variables

| Total path | Rural areas |  | Urban areas |  | Rural vs Urban |  |  |
| --- | --- | --- | --- | --- | --- | --- | --- |
| | $p^{(1)}$ | $se(p^{(1)})$ | $p^{(2)}$ | $se(p^{(2)})$ | $\vert p^{(1)} - p^{(2)}\vert $ | $t$ | $P$ |
| PE→PQ | 0.548 | 0.034 | 0.588 | 0.028 | 0.042 | 0.954 | 0.340 |
| PE→PQ | 0.568 | 0.036 | 0.610 | 0.032 | 0.040 | 0.834 | 0.404 |
| PE→PS | 0.534 | 0.036 | 0.627 | 0.029 | 0.095 | 2.087 | 0.037 |
| PE→PC | -0.114 | 0.016 | -0.166 | 0.020 | 0.052 | 2.034 | 0.042 |
| PE→PT | 0.360 | 0.033 | 0.378 | 0.029 | 0.019 | 0.442 | 0.659 |
| PQ→PV | 0.697 | 0.032 | 0.662 | 0.030 | 0.032 | 0.724 | 0.469 |
| PQ→PS | 0.762 | 0.025 | 0.714 | 0.025 | 0.048 | 1.380 | 0.168 |
| PQ→PC | -0.164 | 0.024 | -0.189 | 0.023 | 0.026 | 0.773 | 0.440 |
| PQ→PT | 0.514 | 0.027 | 0.429 | 0.025 | 0.083 | 2.277 | 0.023 |
| PV→PS | 0.499 | 0.042 | 0.448 | 0.043 | 0.056 | 0.920 | 0.358 |
| PV→PC | -0.107 | 0.018 | -0.119 | 0.017 | 0.011 | 0.442 | 0.659 |
| PV→PT | 0.336 | 0.032 | 0.270 | 0.030 | 0.069 | 1.589 | 0.112 |
| PS→PC | -0.214 | 0.029 | -0.265 | 0.029 | 0.051 | 1.229 | 0.219 |
| PS→PT | 0.674 | 0.028 | 0.602 | 0.029 | 0.071 | 1.741 | 0.082 |

## Discussion

China’s medical reform has entered a critical period, there is an urgent need to understand participants’ satisfaction with BMI and to determine the main aspects that need improvement. This work will provide important evidence for the performance improvement of the insurance schemes and provide a critical reference for reform [4]. After more than 60 years of reform and construction, China has finally established a basic medical insurance system that covers the entire population for basic medicine. To meet participants’ growing basic need for a better life, it is necessary to continuously improve the quality of universal medical insurance from the perspective of the insured [17].

Moreover, the quality of medical insurance determines the satisfaction of the insured to a large extent. The original SIM-URRBMI developed by Peng et al has provide a standard measurement of satisfaction, but could not provide detailed information for the diagnosis of poor quality as there were few detailed quality items. This study constructed the perceived quality variable as a second-order latent variable based on the literature review and expert consultation. The second-order SEM can not only reduce the number of structural model relationships but also make the model path more concise and easier to understand. Most importantly, it can measure the quality perception of URRBMI participants more accurately. This characteristic will greatly help identify the aspects of URRBMI that are in urgent need of improvement [18]. The results of the predictive ability evaluation showed that the 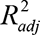 of PS was 0.783, which is larger than the *R*^2^ of PS in the original version of SIM-URRBMI constructed by Peng et al. (0.63). From this point of view, it appears that the construction of PQ as a second-order latent variable in this study increased the predictive power of public satisfaction. This also verifies our hypothesis that the quality-refined model can measure PS more accurately.

Additionally, this study also tested the path effect differences between rural and urban participants; thus, it can not only reveal population heterogeneity in structural models but also help identify the preferred path for quality and satisfaction improvement in a certain subgroup population.

The measurement variables selection is a key step in model construction. CTT and IRT complementary each other, combining both methods in measurement variable selection will provide more stable and reliable choices and benefits for model construction. The result of measurement variable selection revealed that the measurement variable PC2 (formal complaint) had poor performance. It is consistent with Chinese cultural backgrounds that the public seldom choose this method to express their dissatisfaction with public services. Deletion of PC2 suggested that this study had a good rationale for its measurement variable selection methods [19].

The Cronbach’s α coefficients and composite reliability coefficients for all latent variables were greater than 0.7, suggesting good internal consistency. except for the perceived quality variable, which was slightly less than the linear correlation coefficient between the perceived value and public satisfaction variables (One possible reason is that the second-order external factor loadings for the PQ1-PQ3 variables were less than 0.7), all the other results indicate that the model has good convergent validity and discriminant validity. The total effect of perceived quality on public satisfaction is 0.737, which has the largest total effect among all latent variables, indicating that PQ is the most important factor affecting public satisfaction, which is consistent with the research conducted by Lang Xu and Fornell C in the medical insurance field and Johnson MD in other research fields [20-21]. First, from the results of the path coefficients between perceived quality and its first-order latent variables, it was seen that the quality of medical insurance policy had the greatest impact on PQ with a total effect of 0.472, suggesting that medical insurance policy had the greatest impact on public satisfaction. Second, PQ9 (basic coverage scope), PQ10 (coinsurance), PQ11 (deductible) and PQ12 (capitation) had larger second-order factor loadings on PQ. Moreover, PQ9-PQ12 had larger discrimination parameters (2.4-2.8) from the IRT *IIF* results. According to Juxiang Qin’s point of view, the discrimination parameter reflects the sensitivity, which can distinguish the sensitivity of all the participants’ responses to the question (that is, whether the project is a concern shared by everyone), and the *IIF*result reflects that the insured residents are more concerned about the policies related to reimbursement [22]. Therefore, reimbursement policy for URRBMI has the greatest impact on the satisfaction of the insured residents, suggesting that the improvement of quality, especially the medical insurance reimbursement policy quality, is crucial to improving satisfaction with URRBMI.

The SIM-URRBMI satisfaction index score of the main family decision makers for the pupils’ BMI in Changsha was 57.59, which was lower than the satisfaction scores of the insured farmers on the NCMS in 2014 (66.33) and the US federal government satisfaction index scores (69.70) in 2017 [21, 23]. The satisfaction index score can also be used as a vertical comparison tool for the satisfaction research for URRBMI assessment. The satisfaction index scores were 57.29 and 58.17 in the rural and urban areas, respectively, indicating that the urban residents had a slightly higher satisfaction level of URRBMI in Changsha, which is consistent with the research conducted by Fan Yan et al. [24]. The PLS-MGA result showed that, compared to the rural residents, urban participants had a higher total effect of PE on both PS and PC, while having a lower total effect of PQ on PT. PE is a kind of expectation for future medical insurance policies based on past consumption experiences. The urban residents had better policy welfare than the rural residents before the consolidation. Thus, urban residents had higher expectations for URRBMI, and whether the expectation is close or far from the actual situation, they may have, accordingly, had higher positive effects (satisfaction) or negative effects (complaints) than rural residents.

There are also some limitations of this research. First, the sample population was the main family decision makers for pupils’ URRBMI and did not involve the other URRBMI participants in the family (such as college students). The conclusion is limited to the entire insured population. It is hoped that different insured populations can be included in future stages to expand the applicability of the model. Second, this study is a cross-sectional study that did not collect longitudinal data and, thus, could not estimate trends in URRBMI satisfaction in the sample area.

## Conclusions

The final version of the SIM-URRBMI consists of 28 measurement variables and 11 latent variables. It is a reliable and valid satisfaction measurement tool with good prediction ability for public satisfaction and public trust. In addition, the model provides accurate assessment for perceived quality of URRBMI, which will greatly help for performance improvement.

## Data Availability

All data are fully available from the corresponding author by request.

## Acknowledgements

The authors would like to thank all the participants, teachers, and leaders in the selected 8 primary schools, Liuyang City Center For Disease Control and Prevention and Tianxin District Education Bureau.

## Supporting information

**S1 Table.The measurement scores and comparative analysis between two groups of the initial draft of SIM_URRBMI**.

**S2 Table.Item parameter estimation from the GRM.**

**S3 Table.The result of the collinearity assessment in the revised SIM_URRBMI.**

**S4 Table.The indirect effects in the revised SIM_URRBMI.**

### Funding

This work was supported by the project of “13th five year plan” of Hunan Education Science in 2019 (XJK19BGD003), the China Community Health Services & Health Education Program (2014CC03), and education and teaching reform research project of Central South University in 2019 (2019jy146).

## Notes

### Competing Interest Statement

The authors have declared no competing interest.

### Author Declarations

The study protocol was reviewed and approved by the Medical Ethics Committee at Xiang-ya School of Public Health, Central South University. Written informed consent was obtained for each participant who was the medical insurance decision maker of the family.

